# Dissecting Heterogeneity in Functional Network Connectivity Aberrations in Antipsychotic Medication-Naïve First Episode Psychosis Patients – A Normative Modeling Study

**DOI:** 10.1101/2024.08.23.24312480

**Authors:** Rhea A. Thukral, Jose O. Maximo, Adrienne C. Lahti, Saige E. Rutherford, Jordan S. Larson, Hui Zhang, Andre F. Marquand, Nina V. Kraguljac

## Abstract

**Importance:** While there is a general consensus that functional connectome pathology is a key mechanism underlying psychosis spectrum disorders, the literature is plagued with inconsistencies and translation into clinical practice is non-existent. This is perhaps because group-level findings may not be accurate reflections of pathology at the individual patient level.

**Objective:** To characterize inter-individual heterogeneity in functional networks and investigate if normative values can be leveraged to identify biologically less heterogeneous subgroups of patients.

**Design, Setting, and Participants:** We used data collected in a case-control study conducted at the University of Alabama at Birmingham (UAB). We recruited antipsychotic medication-naïve first-episode psychosis patients from UAB outpatient, inpatient, and emergency room settings.

**Main Outcome(s) and Measure(s):** Individual-level patterns of deviations from a normative reference range in resting-state functional networks using the Yeo-17 atlas for parcellations.

**Results:** Statistical analyses included 108 medication-naïve first-episode psychosis patients. We found that there is a high level of inter-individual heterogeneity in resting-state network connectivity deviations from the normative reference range. Interestingly 48% of patients did not have any functional connectivity deviations, and no more than 11.1% of patients shared functional deviations between the same regions of interest. In a *post hoc* analysis, we grouped patients based on deviations into four theoretically possible groups. We discovered that all four groups do exist in our experimental data and showed that subgroups based on deviation profiles were significantly less heterogeneous compared to the overall group (positive deviation group: z= -2.88, p = 0.002; negative deviation group: z= -3.36, p<0.001).

**Conclusions and Relevance:** Our findings experimentally demonstrate that there is a high level of inter-individual heterogeneity in resting-state network pathology in first-episode psychosis patients which support the idea that group-level findings are not accurate reflections of pathology at the individual level. We also demonstrated that normative functional connectivity deviations may have utility for identifying biologically less heterogeneous subgroups of patients, even though they are not distinguishable clinically. Our findings constitute a significant step towards making precision psychiatry a reality, where patients are selected for treatments based on their individual biological characteristics.

**KEY POINTS:** *Question:* How heterogeneous is individual-level resting-state functional network pathology in patients suffering from a first psychotic episode? Can normative reference values in functional network connectivity be leveraged to identify biologically more homogenous subgroups of patients?

*Findings:* We report that functional network pathology is highly heterogeneous, with no more than 11% of patients sharing functional deviations between the same regions of interest.

*Meaning:* Normative modeling is a tool that can map individual neurobiological differences and enables the classification of a clinically heterogenous patient group into subgroups that are neurobiologically less heterogenous.

## INTRODUCTION

Psychosis spectrum disorders are clinically and biologically complex^1^. There is general consensus in the field that functional connectome pathology is central to the underlying pathophysiology^2–4^, but there remains a high degree of unexplained inconsistencies of findings in the existing literature^5,6^. This is perhaps not surprising, given that the knowledge largely stems from case-control studies describing connectome pathology of the ‘average patient,’ while it is becoming increasingly clear that pathological signatures identified at the group-level may not be accurate reflections of pathology at the individual patient level^7–11^.

Normative modeling is a newer statistical approach, inspired by the principles of pediatric growth curve charting, that allows quantification of brain deviations at the individual level^7^ in the context of a normative reference range. Several studies have now demonstrated that inter-individual heterogeneity in brain structures is considerable in schizophrenia, bipolar disorder, autism spectrum disorders, and ADHD, suggesting that group differences are inaccurate reflections of individual-level structural abnormalities^9–12^. More recently, normative reference curves have also been developed for large-scale resting-state brain networks^13^, but to our knowledge, only one study thus far has leveraged this to describe functional brain network pathology at the individual level in schizophrenia^14^. The authors found a high degree of heterogeneity in functional network connectivity disruptions^14^. They also demonstrated that normative data outperformed traditional metrics (e.g. functional connectivity expressed as beta-weights) in disease classification and prediction of cognitive ability in medicated patients with chronic schizophrenia^14^, again, suggesting that individual-level measures outperform traditional metrics of brain function.

Here, we built on the emerging literature and used normative modeling of resting-state brain networks to characterize individual-level pathology in a large group of antipsychotic medication-naïve first-episode psychosis patients, which has not been done before. We hypothesized that functional connectome pathology would show significant inter-individual heterogeneity. We also tested, for the first time, if normative deviations have potential utility in identifying more biologically homogeneous subgroups of patients. The ability to identify groups of patients based on shared biological features constitutes a critical step towards precision medicine, where treatments selected based on biological features.

## METHODS

Patients were recruited through the University of Alabama at Birmingham (UAB) Emergency Department, inpatient, and outpatient units. Ethical approval was obtained through the UAB Institutional Review Board (IRB), and informed consent was obtained from all participants (participant assent and parental consent was obtained for participants younger than 18). We also assessed decisional capacity for informed consent in patients^15^. Healthy volunteers were recruited through flyers and advertisements posted throughout the Birmingham area.

Healthy controls and patients were excluded if they had a central nervous system illness, any major medical conditions, history of head trauma with loss of consciousness >2 minutes, MRI contraindications such as metal objects, an active moderate to severe substance use disorder within the past month (excluding nicotine and cannabis), or if they were pregnant. Patients were further excluded if they had more than five days lifetime exposure to antipsychotic medications. Healthy controls were further excluded if they had a psychiatric history or a first degree relative having a psychotic disorder.

Consensus diagnoses were made according to DSM-5 criteria by two board-certified psychiatrists (ACL and NVK), taking into consideration information from the Diagnostic Interview for Genetic Studies (DIGS)^16^ or Mini-International Neuropsychiatric Interview 7.0 (MINI)^17^ and from all historical and direct assessment information available. The Brief Psychiatric Rating Scale (BPRS)^18^ and Repeatable Battery for the Assessment of Neuropsychological Status (RBANS)^19^ were used to assess clinical symptom severity and cognition, respectively.

### Data acquisition

All imaging data were acquired using a 3T whole-body Siemens MAGNETOM Prisma MRI scanner equipped with a 20-channel head-coil. A high-resolution T1-weighted structural scan was acquired for anatomical reference (MPRAGE: TR/TE/TI= 2400/ 2.22/1000ms; flip angle = 8°; GRAPPA factor = 2; voxel size = 0.8x0.8x0.8mm^3^; FOV = 256x256mm). Two resting-state functional MRI (rs-fMRI) runs were acquired in opposing phase encoding directions (anterior > posterior and posterior > anterior; TR/TE= 1550/ 37.80ms; flip angle = 71°, multi-band acceleration factor = 4; voxel size= 2x2x2mm^3^; 225 volumes, and 72 axial slices; FOV = 208x208mm). During the resting state scan, participants were presented with a fixation cross and instructed to keep their eyes open and let their mind wander.

### Image processing

T1-weighted images were skull-stripped using FSL BET^20^. The first 10 volumes of each fMRI run were removed to allow for signal equilibration and then corrected for magnetic field inhomogeneities and integrated using FSL topup^21,22^. Functional images were then imported into the CONN toolbox (version 21a)^23–25^ where they were corrected for slice timing and motion using rigid-body alignment, co-registered to the skull-stripped structural image, normalized to Montreal Neurological Institute (MNI) space to allow for later parcellation using the Yeo-17 atlas^26^, bandpass filtered (0.008 Hz – 0.08 Hz), and spatially smoothed with a 4-mm full width at half maximum (FWHM) Gaussian kernel. Outlier volumes were censored using the artifact detection (ART) toolbox^23,24^ (composite volume-to-volume motion >0.9 mm and global signal intensity >5SDs). Framewise displacement and percentage of censored data were then calculated to determine if scans should be excluded from analysis; scans with a mean framewise displacement >2mm or had >50% of censored data were excluded. The six motion parameters derived from rigid-body realignment and their derivatives, as well as the first component time series derived from cerebrospinal fluid and white matter using aCompCor^23^ and corresponding derivatives, were regressed out from the signal. Global signal regression was not performed as this processing step has been shown to introduce false negative correlations^27^. Functional brain networks were then parcellated using the Yeo-17 functional network parcellation atlas^26^, resulting in a functional connectivity matrix of 136 connectivity values.

During post-processing quality control, if processed resting-state fMRI images were found to not have correctly co-registered to the structural image or normalized to the MNI space, the registration algorithm, Advanced Normalization Tools (ANTs)^28^, was used to correct the registration mistakes.

### Normative Modeling

Here, we used the Predictive Clinical Neuroscience (PCN) toolkit (version 0.27), a publicly available Python package, for normative modeling of resting state connectivity data. The normative model was generated using resting state connectivity data from approximately 22,000 healthy individuals across the lifespan (age range (yrs): 2-100). Healthy control data collected in this project were used for calibration of the normative reference curves (Python version 3.8.3^29,30^, PCN toolkit version 0.27^31^).

### Statistical analyses

Statistical analyses and plot generation were conducted using R version 4.3.1^32^. We quantified functional connection deviations expressed as standard deviations (z-score) from the normative reference range for each patient. We binarized z-scores to operationally define a significant deviation using a cut-off value of ±2 standard deviations. We then calculated connectivity deviations between the functional connections and calculated the deviation load (number of absolute, positive, and negative deviations in an individual patient), and the percentage of participants who share the same functional connection deviations (i.e. functional connection deviation similarities between the same regions of interest). To evaluate the distribution of the data, we created a scatter plot of individual-level deviation load data across two axes (x = negative deviation load, y = positive deviation load).

In *post hoc* analyses, we examined if network deviation profiles allow us to identify biologically less heterogenous subgroups. Given that group-level studies have been inconsistent and reported patterns of only increased connectivity^2,6^, only decreased connectivity^33,34^, or a mix of increased and decreased connectivity^35,36^ in psychosis spectrum disorders, we made the decision to examine individual-level deviation patterns in the same categorical framework. We grouped individuals according to plausible functional deviation profiles: (1) patients that have only positive deviations (increased connectivity), (2) patients that have only negative deviations (decreased connectivity), (3) patients that have a mix of positive and negative deviations, and (4) patients that display no significant functional connectivity deviations. We then mapped connectivity deviation patterns for each group separately, correcting for multiple comparisons by controlling for the false discovery rate [α= 0.05 significance threshold]. To do so, we set the significance threshold for the total number of positive/ negative deviations 4 or greater [number of connections (136) x significant one-tailed alpha (0.025) = 3.4 (4 upon rounding)]. To determine if the functional connection deviations with the highest patient similarity percentage within each subgroup were less heterogeneous than the maximum overall patient group similarity percentage, a z-test was used.

## RESULTS

A total of 108 antipsychotic medication-naïve first-episode psychosis patients were included in the analyses. For demographic and clinical characteristics of the patient and healthy control groups, refer to Table 1.

**Table 1:** Demographic and Clinical Characteristics.

|  | Patients (N = 108) | HC (N = 122) | Test | P value |
| --- | --- | --- | --- | --- |
| Age, mean(SD), y | 23.54(5.49) | 23.59(5.10) | t = 0.07 | 0.94 |
| Sex |  |  |  |  |
| Male | 65 | 70 | $\chi^2 = 0.09$ | 0.77 |
| Female | 43 | 52 |  |  |
| Socioeconomic Status, mean(SD) | 5.69(4.94) | 4.58(4.10) | t = 1.81 | 0.07 |
| Packs Per Day, mean(SD), day | 0.23(0.39) | 0.01(0.06) | t = 5.72 | <0.001 |
| Cannabis Use(% positive) | 41.67 | 1.6 | $\chi^2 = 52.90$ | <0.001 |
| DUP, mean(SD), months | 22.64(38.69) | NA | NA | NA |
| Diagnosis |  |  |  |  |
| Schizophrenia | 51 | NA | NA | NA |
| Schizoaffective Disorder | 21 | NA |  |  |
| Bipolar Disorder | 4 | NA |  |  |
| Schizophreniform | 7 | NA |  |  |
| Psychosis NOS | 18 | NA |  |  |
| Brief Psychotic | 5 | NA |  |  |
| MDD with Psychosis | 2 | NA |  |  |
| BPRS |  |  |  |  |
| Positive 4, mean(SD) | 14.81(3.86) | NA | NA | NA |
| Negative, mean(SD) | 5.56(3.12) | NA |  |  |
| Total, mean(SD) | 48.28(11.12) | NA |  |  |
| RBANS |  |  |  |  |
| Immediate Memory, mean(SD) | 83.60(18.03) | 103.3(14.05) | t = -8.56 | <0.001 |
| Visuospatial, mean(SD) | 75.84(17.13) | 83.73(13.27) | t = -3.61 | <0.001 |
| Language, mean(SD) | 85.08(16.74) | 97.3(14.86) | t = -5.45 | <0.001 |
| Attention, mean(SD) | 81.74(18.01) | 102.5(14.66) | t = -8.87 | <0.001 |
| Delayed Memory, mean(SD) | 77.08(15.89) | 91.75(10.37) | t = -7.61 | <0.001 |
| Total, mean(SD) | 75.45(15.91) | 93.75(10.62) | t = -9.43 | <0.001 |
| Race |  |  |  |  |
| African American | 63 | 25 | NA | NA |
| Asian | 4 | 23 |  |  |
| Caucasian | 39 | 66 |  |  |
| Hispanic/Latino | 1 | 4 |  |  |
| Other | 1 | 4 |  |  |
| Ethnicity |  |  |  |  |
| Not Latino/Hispanic | 106 | 117 | NA | NA |
| Latino/Hispanic | 2 | 5 |  |  |
| Not Answered | - | - |  |  |

Network perturbations were highly heterogeneous between individual patients, with no more than 11.1% of patients sharing positive deviations in the same network connection and no more than 8.3% of patients sharing negative deviations in the same network connection (Figure 2B).

**Figure 1:**
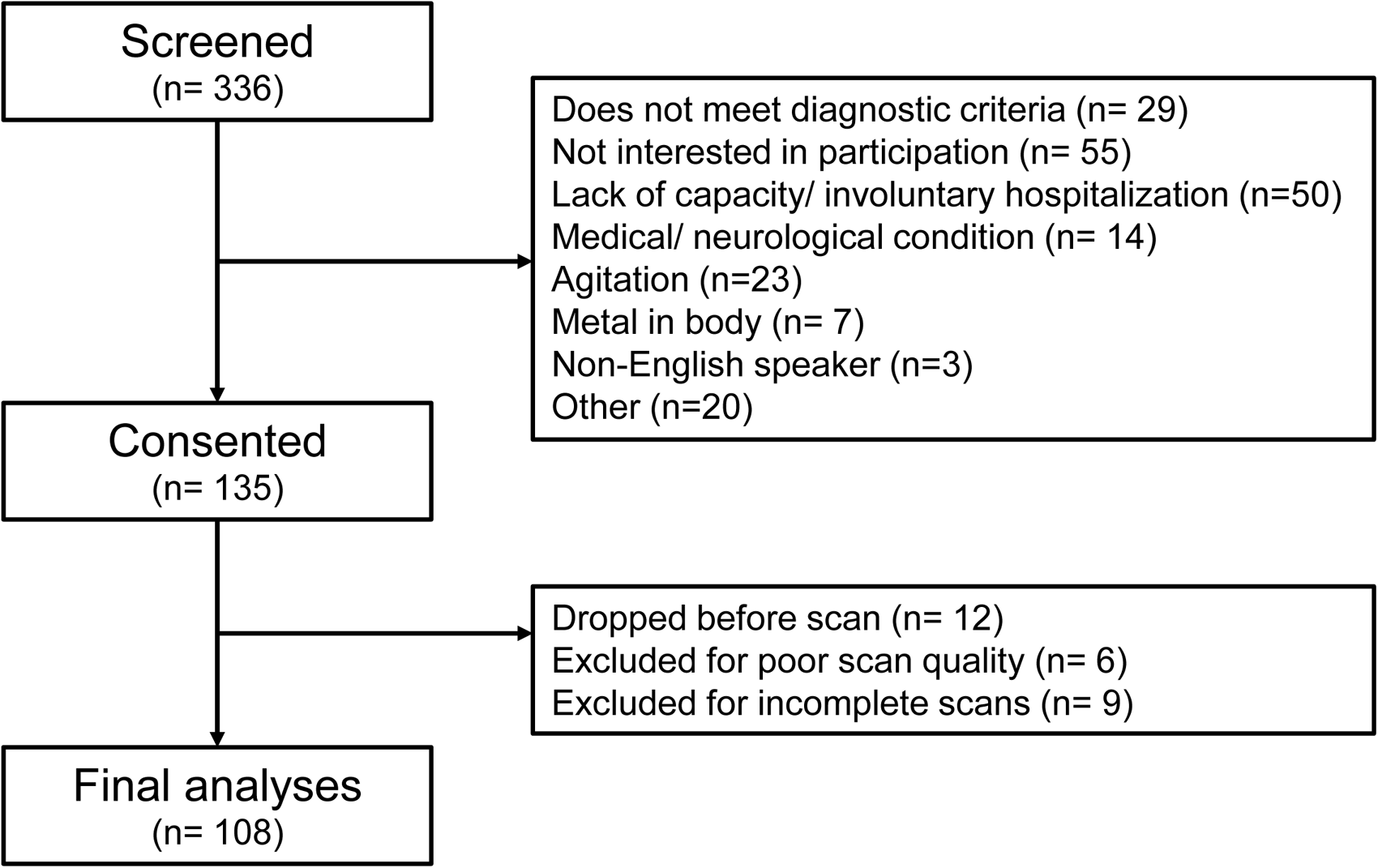
Consort chart. 336 patients were screened, of which 201 were excluded prior to consent, 135 patients were consented and 108 patients were included in final analyses.

**Figure 2:**
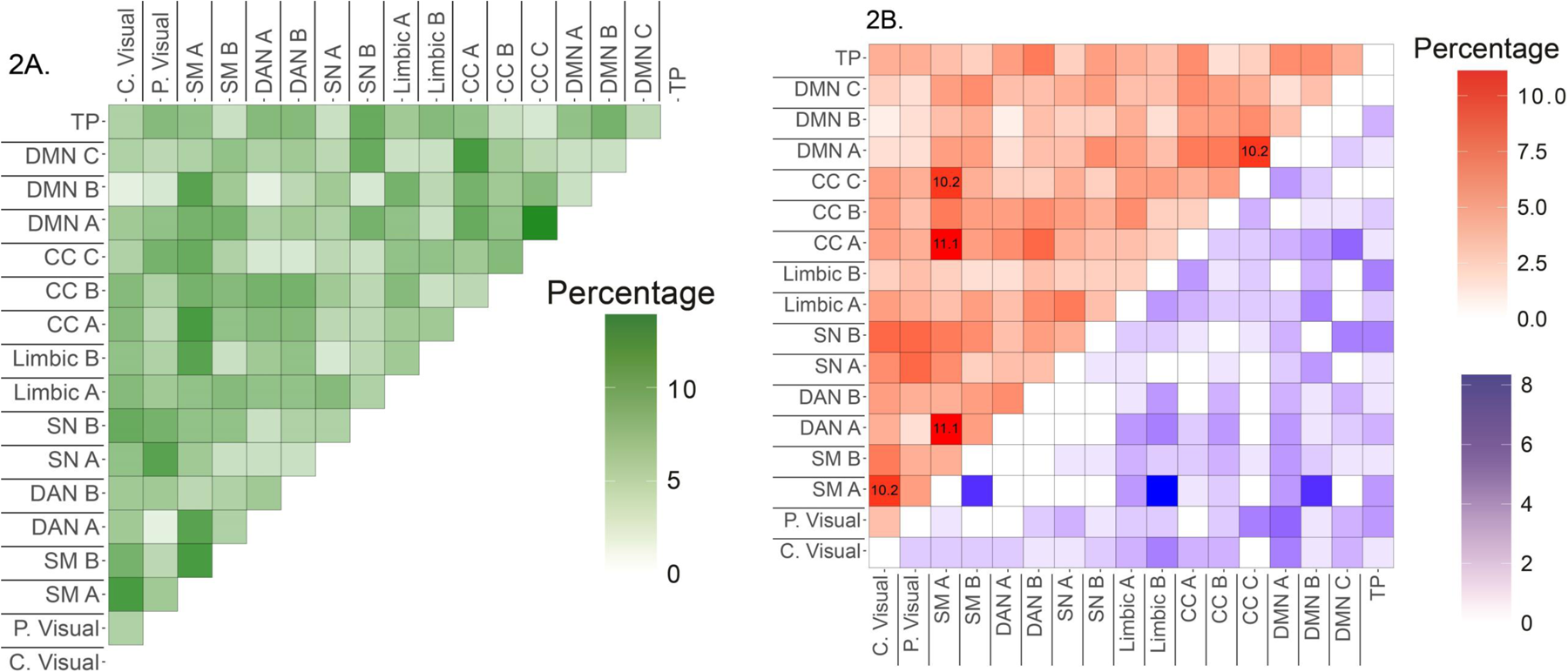
Heterogeneity in absolute, positive, and negative functional network connectivity deviations. 2a. Percentage of patients who share absolute functional network connectivity deviations (≥ |2| standard deviations) within a network. Figure 2b. Percentage of patients who share a positive (≥ 2 standard deviations) or negative (≤ -2 standard deviations) deviation within a network. Percent of positive deviations are in red, negative deviations in blue. Functional network deviations that are shared by ≥ 10% of patients include an indication of the percentage value in the heatmap. C. Visual = Central Visual; P. Visual = Peripheral Visual; SM A = Somatomotor A; SM B = Somatomotor B; DAN A = Dorsal Attention Network A; DAN B = Dorsal Attention Network B; SN A = Salience/Ventral Attention Network A; SN B = Salience/Ventral Attention Network B; CC A = Cognitive Control Network A; CC B = Cognitive Control Network B; CC C = Cognitive Control Network C; DMN A = Default Mode Network A; DMN B = Default Mode Network B; DMN C = Default Mode Network C; TP = Temporal Parietal.

In *post hoc* analyses, we grouped patients according to plausible subgroups (positive only, negative only, mixed, none) and discovered that all four subgroups do exist in experimental data. Approximately 52% of patients had a significant number of connection deviations, whereas 48% patients had no significant functional connection abnormalities (Figure 3). Of the 52%, approximately 25% of patients had exclusively positive connectivity deviations, 24% of patients had exclusively negative connectivity deviations, but only 2.8% had a mix of both positive and negative deviations (Figure 3). In addition, the spatial patterns of deviations within the subgroups were less heterogeneous compared to the overall group. In the positive only deviation group, 40.7% of patients shared deviations in the same functional connections (z= - 3.66; p=0.0001). The most consistently shared deviations in the group were in SM A network connectivity to DAN-A, CCN-C and C. Visual network, as well as DMN-A and CCN C networks. The negative only deviation group showed up to 23.1% in deviation similarities, which is more than twice as common as in the overall group (z= -2.15, p= 0.016). Deviations in this group were most commonly shared in connections of the SM A and Limbic A network (refer to Figure 2 description for the abbreviation legend). Given that only 3 patients fell into the category of mixed deviations, we did not have sufficient power to make meaningful interpretations regarding heterogeneity in that group (all subgroup data are illustrated in Figure 4).

**Figure 3:**
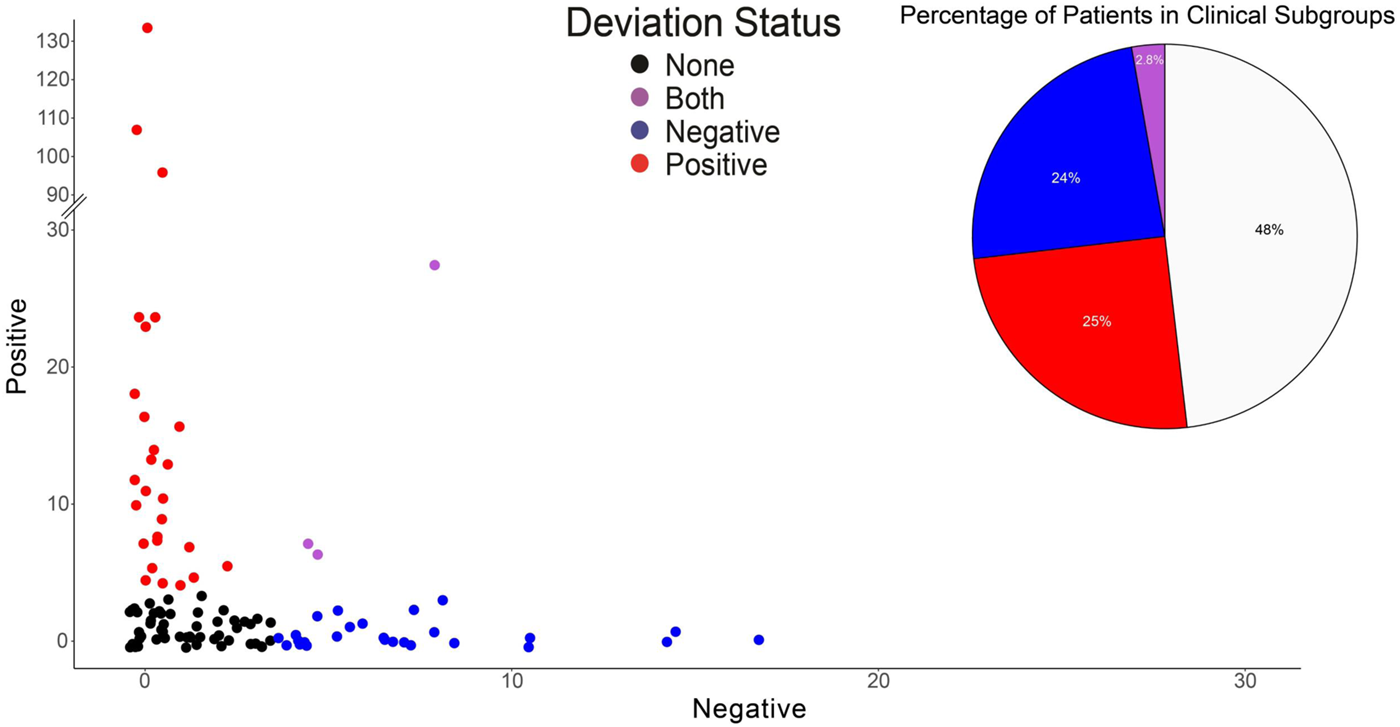
Deviation Load Distributions. Positive and negative deviation loads for each patient plotted as a scatterplot. Black: none group, red: mostly positive group, blue: mostly negative group, purple: mixed group.

**Figure 4:**
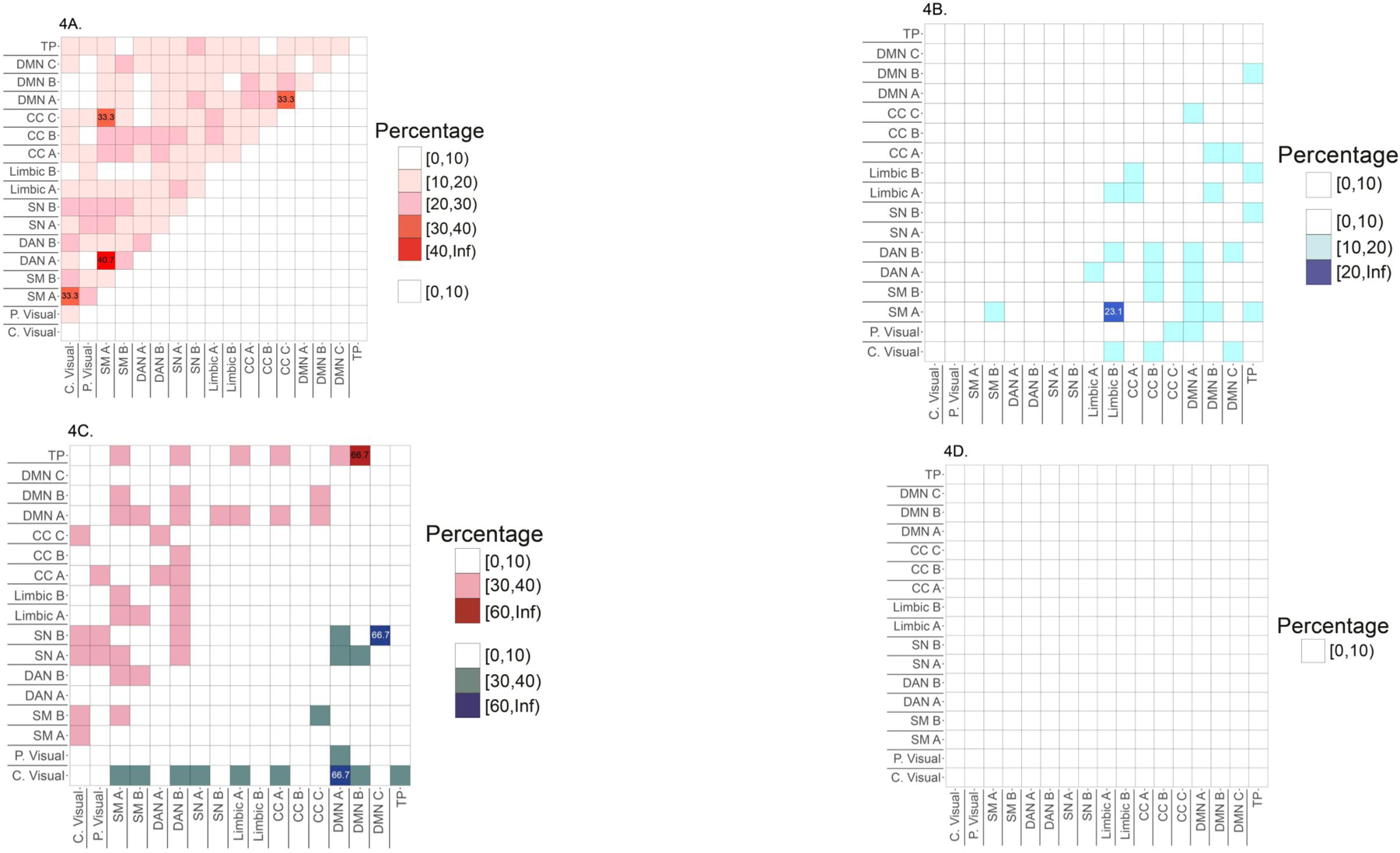
Functional connectivity deviations in patients grouped based on deviation loads. 4a. Percent of patients in the group who share functional network connectivity deviations in the mostly positive group (N = 27), 4b. Percent of patients in the group who share functional network connectivity deviations in the mostly negative group (N = 26), 4c. Percent of patients in the group who share functional network connectivity deviations in the mixed positive/ negative deviation group (N = 3), 4d. Percent of patients in the group who share functional network connectivity deviations in the no deviation group (N = 52). Left half of heatmaps map percentage of positive deviation similarities, right half maps percentage of negative deviation similarities.

Patients in these subgroups did not differ in terms of clinical presentation (BPRS total, positive, and negative symptom scores) or RBANS scores (language, attention, delayed memory, immediate memory, and visuospatial scores) (Supplemental Table 1). Furthermore, cannabis use was not significantly different between the four subgroups (χ^2^ = 3.8, p = 0.31) (Supplemental Table 1).

## DISCUSSION

To our knowledge, this is the first resting-state connectivity study using normative modeling to quantify aberrant brain network connectivity at the individual level in medication-naïve first-episode psychosis patients. Here, we present two findings highlighting that group-level analyses are inaccurate representations of individual-level disease pathology: (1) Functional connection deviations are heterogeneous across individuals, where no more than 11% of patients overall share the same affected functional connections, and (2) approximately 48% of first-episode patients did not have a significant number of functional connection deviations. Perhaps most importantly, we found evidence that all four plausible subgroups exist in experimental data, and that subgroups were less heterogeneous in deviation patterns. Our findings experimentally support the idea that pathophysiological findings in psychosis spectrum disorders are heterogeneous, and suggest the evidence of biologically less heterogeneous subgroups. This marks an important milestone for moving closer to making precision psychiatry a reality, where patients will be selected for treatments based on their biological characteristics.

Contradictory results across connectome studies, with reports of hyper-connectivity^2,6^, hypo-connectivity ^33,34^, or mixed functional network connectivity abnormalities^35,36^ in psychosis spectrum disorders, are typically attributed to differences in patient populations such as illness stage^36^, medication status^37^, image acquisition parameters, or data-analysis techniques. While these factors are important sources of variability, our data suggests that inter-individual variations in functional connectome pathology may be a key source of inconsistencies in the literature. Our findings are consistent with two studies in chronic schizophrenia that demonstrated a high level of heterogeneity in functional connectivity networks^14,38^. The prevailing hypothesis for the presence of individual-level differences is that multiple disease mechanisms converge on a clinical phenotype, and that subgroups of patients are expected to share a common causal mechanism^39^, which is also supported by our data.

Interestingly, we found experimental evidence of all four plausible subgroups defined based on deviation loads. First, we would like to highlight a rather unexpected finding, namely that 48% of patients did not show any significant deviations in their functional connectome compared to the normative reference range. Importantly, clinical disease expression did not differ between subgroups, so this finding cannot be explained by low symptom burden in this group (Supplementary Table 1). These results challenge the conventional notion of functional connectome pathology as a core feature of schizophrenia and underscore the importance of characterizing disease pathology at the individual level. To our knowledge, only a few reports in the literature have explicitly highlighted the existence of a subgroup of patients with a psychosis spectrum disorder that appear neurobiologically intact. For example, Chand and colleagues described two distinct neuroanatomical subtypes in chronic, medicated patients with schizophrenia, where one group was characterized by widespread cortical gray matter volume loss, while the other subtype (approximately 37% of the sample) did not show any cortical thickness abnormalities^40^, only basal ganglia volume increase, which could be explained by antipsychotic medication effects. Pan and colleagues identified a ‘healthy-like, morphologically intact’ subgroup of early-stage schizophrenia patients using a cluster analysis of cortical thickness^41^. Similarly, another group using cluster analyses showed that one of three biological subtypes of schizophrenia had normal cortical and subcortical volumes, interestingly, this was present both in first-episode psychosis patients and chronic schizophrenia patients^42^. Our findings add to the emerging literature suggesting that a subtype in schizophrenia is characterized by the lack of structural abnormalities, by demonstrating, for the first time, that there is also a group of patients with psychosis spectrum disorders that has no apparent functional network connectivity abnormalities. However, it is important to note that our investigation only included cortical functional networks. It remains unknown if this subtype is characterized by subcortical rather than cortical functional connectivity pathology.

While we hypothesized that we would detect biologically more similar subgroups, we found it curious to realize that most patients who had connectivity deviations, had deviations exclusively in either the positive or negative direction; only about 3% of patients had a mix of positive and negative deviations. The positive deviation group most commonly shared hyperconnectivity between the primary motor (SM-A) and visual cortices (C. Visual), SM-A and the DAN-A, the SMA-A and CCN-A, and the CCN-C and DMN-A. In contrast, the negative deviation group most commonly shared hypoconnectivity between the SM-A and the limbic B network. While both groups converge on primary motor functional deviations, the positive deviation group most commonly shares functional network pathology between the SM-A and higher-cognitive networks in the neocortex, whereas the negative deviation group most commonly shares functional network pathology between the SM-A and the limbic cortex, which is phylogenetically older and consists of four rather than six cortical layers. Together, these data show that the four subgroups, albeit converging on a clinical phenotype, are characterized by different disease signatures. It also demonstrates that normative modeling may have utility in identifying biologically less heterogeneous subtypes of the disorder, which could eventually be leveraged for patient selection in precision psychiatry treatments.

One of the major strengths in this study is that we enrolled a large group of antipsychotic medication-naïve first-episode psychosis patients, which allows us to mitigate confounds of illness chronicity and antipsychotic medication exposure^39^. We chose not to exclude patients who used cannabis, a major risk factor for developing psychosis, as this would have limited the generalizability of our data. The normative models used here likely overrepresent individuals with European ancestry, which do not match our patient demographics. Prior studies suggest that transferring the model to diverse data should be done with caution. We acknowledge this as a major limitation given that many of the patients in this study identify (63.89%) as a racial/ ethnic minority, a group that has historically lacked equitable inclusion in research studies.

In summary, our data supports the idea that group-level findings are not an accurate representation of functional connectome pathology at the individual patient level. Furthermore, characterizing individual-level pathology shows promise for identification of biologically less heterogeneous subgroups of patients, which is a foundational step to identification of new treatment targets and precision medicine approaches. As immediate next steps, it will be important to independently replicate this finding in a different group of first-episode psychosis patients and to determine how stable these subtypes are as the illness progresses. It will also be critical to better understand individual-level pathology across multiple levels of analysis which has the potential to shed light on the intricate interplay between structural, microstructural, and functional pathology in psychosis spectrum disorders.

## Data Availability

All data produced in the present work are contained in the manuscript.

---

**Supplemental Table 1:** Demographic and Clinical Characteristics of the Four Subgroups.

|  | Positive (N = 27) | Negative (N = 26) | Both (N = 3) | None (N = 52) | Test | P value |
| --- | --- | --- | --- | --- | --- | --- |
| Age, mean(SD), y | 21.78 (3.51) | 23.15 (5.75) | 27.38 (4.04) | 24.63 (6.10) | F=1.69 | 0.17 |
| Sex |  |  |  |  |  |  |
| Male | 19 | 18 | 1 | 27 | X<sup>2</sup>=4.44 | 0.23 |
| Female | 8 | 8 | 2 | 25 |  |  |
| Socioeconomic Status, mean(SD) | 5.37 (4.72) | 5.15 (4.70) | 4.67 (5.11) | 6.20 (5.32) | F=0.35 | 0.79 |
| Packs Per Day, mean(SD), day | 0.25 (0.49) | 0.14 (0.30) | 0.50 (0.5) | 0.25 (0.37) | F=0.96 | 0.42 |
| Cannabis Use (% positive) | 29.63 | 53.85 | 33.3 | 42.31 | X<sup>2</sup> = 3.8 | 0.31 |
| DUP, mean(SD), months | 26.14 (43.56) | 14.52 (26.08) | 1.08 (1.23) | 26.12 (41.94) | F=0.90 | 0.44 |
| Diagnosis |  |  |  |  |  |  |
| Schizophrenia | 11 | 12 | 0 | 28 | NA | NA |
| Schizoaffective Disorder | 7 | 5 | 1 | 8 |  |  |
| Bipolar Disorder | 2 | 0 | 1 | 1 |  |  |
| Schizophreniform | 2 | 1 | 0 | 4 |  |  |
| Psychosis NOS | 5 | 7 | 0 | 6 |  |  |
| Brief Psychotic Disorder | 0 | 1 | 0 | 4 |  |  |
| MDD with Psychosis | 0 | 0 | 1 | 1 |  |  |
| BPRS |  |  |  |  |  |  |
| Positive, mean(SD) | 14.08 (3.29) | 14.65(3.55) | 13.00(6.43) | 15.21 (4.16) | F=0.56 | 0.64 |
| Negative, mean(SD) | 5.92 (2.98) | 5.39(3.14) | 4.00 (1.00) | 5.56 (3.29) | F=0.39 | 0.76 |
| Total, mean(SD) | 49.81 (10.19) | 47.46(9.86) | 44.33 (12.01) | 48.15 (12.25) | F=0.33 | 0.80 |
| RBANS |  |  |  |  |  |  |
| Immediate Memory, mean(SD) | 83.74 (19.11) | 81.7 (14.51) | 92 (2.83) | 84.14 (19.68) | F=0.24 | 0.87 |
| Visuospatial, mean(SD) | 77.39 (13.58) | 76.26 (16.47) | 94 (14.14) | 73.98 (19.04) | F=0.99 | 0.40 |
| Language, mean(SD) | 86.91 (18.42) | 79.0 (19.43) | 91.0 (18.38) | 87.02 (13.84) | F=1.40 | 0.25 |
| Attention, mean(SD) | 79.96 (19.16) | 75.7 (15.78) | 92.50 (2.12) | 85.34 (18.20) | F=1.81 | 0.15 |
| Delayed Memory, mean(SD) | 76.65 (16.07) | 78.65 (13.33) | 83.5 (0.71) | 76.18 (17.53) | F=0.23 | 0.88 |
| Total, mean(SD) | 75.83 (16.84) | 72.3 (14.21) | 87.50 (9.19) | 76.34 (16.51) | F=0.73 | 0.54 |
| Race |  |  |  |  |  |  |
| African American | 12 | 19 | 1 | 31 | NA | NA |
| Asian | 0 | 1 | 0 | 3 |  |  |
| Caucasian | 14 | 6 | 2 | 17 |  |  |
| Hispanic/Latino | 1 | 0 | 0 | 0 |  |  |
| Other | 0 | 0 | 0 | 1 |  |  |
| Ethnicity |  |  |  |  |  |  |
| Not Latino/Hispanic | 25 | 26 | 3 | 52 | NA | NA |
| Latino/Hispanic | 2 | 0 | 0 | 0 |  |  |

## ACKNOWLEDGEMENTS

This work was supported by the National Institute of Mental Health (grant numbers: R01MH118484 [NVK] and R01MH113800 [ACL]).

